# Measuring Depression, and Its Association with Substance Use, during an Adherence Study of Patients Treated for Tuberculosis in Haitian Prisons

**DOI:** 10.1101/2023.09.20.23295740

**Authors:** Lovette C. Ekwebelem, Margarette R. Bury, Edwin J. Prophete, Karine Duverger, Patrick M. Bircher, Anne C. Spaulding

## Abstract

**Purpose:** Diagnosed depression is prevalent in prisons of affluent countries; literature on depression screening in prisons of low-resource nations is sparse. Haiti has experienced multiple recent disasters, which could have both somatic and mental health consequences. To surveil its prisons for depression, ethnoculturally appropriate scales could be helpful.

**Design/methodology/approach:** We performed a cross-sectional analysis of symptoms of depression and its associations among participants in a 2019-2020 tuberculosis treatment adherence project across 6 Haitian prisons. To measure depression, we piloted the use of the Zanmi-Lasante Depression Symptom Inventory (ZLDSI) scale in a carceral setting. We calculated its Cronbach alpha in this setting and generated binary logistic models to study the associations of depression with basic demographic variables; use of cigarettes, marijuana, and alcohol; and incarceration history. We then performed a multivariate logistic regression to determine if substance use and education predicted depression, after adjusting for age.

**Findings:** Fifty subjects were recruited; age ranged from 18 to 59 years. Adherence to TB medication was recorded as above 99% in all subjects. The Cronbach alpha score for the ZLDSI scale in this population was 0.77, signifying the good fit of the scale for this population. A ZLDSI score ≥13.0 has been associated with depression; 66% of participants had scores of 13.0 or greater, mean 13.9 (S.D. 8.2). Multivariate analysis showed significant associations between depression, alcohol consumption, age, and income.

**Originality/Conclusion:** We believe this study represents the first measurement of depressive symptoms in a Haitian prison population; it found symptoms common.

## Introduction

In the literature on morbidity among incarcerated populations in high resource countries, serious mental illness and behavioral issues figure prominently. In contrast, the information on mental health among imprisoned people in low resource countries is sparse. Depression and other mental illnesses are universal health conditions affecting people in all countries of the world regardless of socioeconomic status. Therefore, the absence of literature on the mental health of persons in custody in low-income countries represents a gap in knowledge. Importantly, mental illness may influence adherence to medical regimens and should be addressed when treating other co-morbidities, such as infectious diseases.

Depression is expressed in culturally specific ways (Cénat et al., 2020). To better understand mental health on a global level, one must understand and respect the vast cultural differences among nations. A single, standardized diagnostic scale cannot capture an accurate depiction of mental health in all global environments without considering cultural context. Consequently, organizations have worked to measure mental health using scales that are unique to their own country’s expression of mental distress. In 2014, Partners in Health recognized this need and developed the Zanmi-Lasante Depression Inventory Scale (ZLDSI) for use among Haitian community members ((Rasmussen et al., 2014); See Appendix *Tables S1 and S2*.) They validated the scale with a convenience sample of persons with and without clinician diagnosed depression.

We used this scale when assessing factors leading to adherence to TB therapy in a previously described trial of Video Directed Observed Treatment (VDOT) for TB using the SureAdhere (San Diego CA) platform (de Groot et al., 2022, Kehus et al., 2021). In essence, VDOT was implemented in 5 remote Haitian prisons that had irregular staffing of healthcare workers, as a means to observe TB treatment for more of the country’s prison population. Correctional officers facilitated bringing persons under treatment for TB in front of an electronic tablet at these 5 prisons, which could asynchronously transmit videos via the internet to a study office located in Haiti’s capital, Port-au-Prince. For patients leaving prison before treatment completion, a mobile phone was given to continue to record via the SureAdhere platform that medications were taken. Three prisons fully staffed with healthcare workers served as control prisons, where TB treatment was given via Direct Observed Therapy (DOT).

Substance use is not an insubstantial problem in Caribbean countries. Studies among survivors of the Haiti earthquake in 2010 have discovered increased prevalence of mental health issues along with increased alcohol consumption (Cénat et al., 2020). Other research shows that there may be higher substance use prevalence among those incarcerated in LMICs compared to HICs (Hill et al., 2022), but an association between substance use and incarceration in Haitian populations has not been studied. With mental distress and substance abuse as major problems in Haiti after the 2010 earthquake, more research is needed on measuring substance use and mental health in incarcerated populations in Haiti, and characterizing whether there is correlation.

## Methods

### Survey

All data come from a survey administered in conjunction with a study of correctional officer-facilitated VDOT for TB treatment in Haitian Correctional Facilities that started in 2019 and concluded in at staggered times between April and July of 2020. The survey was conducted at baseline to measure potential predictors of medication adherence, including basic demographics, depression, substance use, and stigma. Items about substance abuse, tuberculosis knowledge, taking medications and demographics came from the initial survey used when testing the SureAdhere methodology in the US (Garfein et al., 2015, Garfein et al., 2018), contextualized for Haiti. For the measurement of stigma about being infected with TB, a scale from Coreil *et al*. piloted in Haiti was used as a baseline, with 7 of 22 items deemed not appropriate for the prison setting and removed, and others modified (See Appendix *Tables S3-S5*). Along with the ZLDSI scale, items were taken from scales in English. English language items were translated into Haitian Creole. The entire survey instrument was then back translated into English.

### Adherence

adherence was measured by nurses in the fully staffed, control prisons during the study period. Data on medication adherence was recorded via video at the intervention prisons using the SureAdhere platform. Per protocol of the National TB Control Program of Haiti, a paper-based log of doses taken was also maintained at the intervention as well as the control prisons (Charles et al., 2017). We reviewed an Excel file that replicated the paper-based form; we also reviewed the SureAdhere logs.

### Subjects and setting

In total, 87 persons were enrolled in the VDOT program during the study period and 102 were enrolled in TB treatment via DOT at the control prisons. All persons in a project prison on a day when the study team visited who were taking TB therapy, regardless for how long, were approached and asked if they wanted to take the associated survey. Participants were men who resided in one of five VDOT intervention prisons (Mirebalais, Petit Goave, Jacmel, Carrefour, and Gonaives). One of these prisons (Carrefour) ended up not fully implementing the VDOT protocol due to a lack of trained correctional officers. One of the control prisons (Croix de Bouquet) using health worker-administered DOT was also included.

### Data Collection

An informed consent procedure was conducted in Haitian Creole. Survey data were collected from 50 participants with TB, aged 18 to 59 years old from 6 Haitian prisons for TB medication adherence. The survey was conducted at all sites by Haitian Creole-speaking researchers reading the questions, and answers were recorded with pen and paper. After data collection, all data were transcribed into REDCap (Research Electronic Data Capture, Vanderbilt University, Nashville, TN).

#### Data Analysis

Descriptive analyses were conducted to observe the overall means and standard deviations (SD) for continuous variables, and totals (n) and percentages (%) for categorical variables. Demographic characteristics of the study participants, measurements of stigma, and use of substances were compared to the probability of a score on the ZLDSI depression scale of 13 or greater - the score used as the cut-off for screening for depression, with balanced sensitivity and specificity, during development and validation of the ZLDSI scale (Rasmussen et al., 2014).

Simple linear regression was performed to study any significant correlations between ZLDSI scores (as a continuous variable) and variables in the study. Bivariate logistic regression was used to analyze any associations between marijuana use, smoking use, alcohol use, age, education, financial income, whether current incarceration is the first instance of confinement, DOT vs VDOT site, stigma associated with depression, number of dependents, and number of hours worked per week prior to incarceration. After examining significance and conducting a collinearity assessment, only alcohol use, age, education, financial income, and depression were kept in a final model. Financial income was kept as a confounder in this model and alcohol use was a mediator. The likelihood ratio test and Wald test were utilized to help build the model. Missing data were omitted from the multivariate logistic analyses.

#### Bioethics Oversight

The study protocol was approved by the Institutional Review Board of Emory University and the Haitian Ministry of Health’s National Bioethics Committee.

## Results

### Baseline Characteristics of Participants by VDOT and DOT Sites

Descriptive statistics of the study population are summarized in *Table 1*. Out of all 50 participants in the study, 39 (78%) were in sites utilizing the VDOT intervention, and 11 (22%) were in one of the control sites using the standard of treatment, DOT. All participants identified as men in this study, which was set exclusively in male Haitian prisons. The average age in the VDOT sites was 31.03 years (SD=8.95) compared to 30.2 (SD=6.44) in the DOT control site surveyed. As for substance use among the overall study population, more participants in total reported alcohol consumption [28(56%)] compared to cigarette use [21(42%)] or marijuana use [11(22%)].

**Table 1.** Characteristics of Study Subjects: Incarcerated TB patients, Haiti, 2019-2020.

| Variable | VDOT sites<br>(Intervention)<br>n = 39 | DOT sites<br>(Control)<br>n = 11 | Overall<br>n = 50 |
| --- | --- | --- | --- |
| **Gender, Male [n(%)]** | 39 (100%) | 11 (100%) | 50 (100) |
| **Age, years [mean (SD)]** | 31.03 (9) | 30.2 (6.4) | 30.8 (8.4) |
| **Attended School [n (%)]** | 23 (59) | 9 (81.8) | 32 (64) |
| *Missing [n (%)]\*\** | 9 (23.1) | 1 (9.1) | 10 (20) |
| **Mean Number of Years Attended**<br>**among Those with Schooling [mean**<br>**(SD)]** | 3.9 (3.7) | 6.7 (4.4) | 4.60 (4) |
| **First Time Incarcerated [n (%)]** | 31 (79.5) | 8 (72.7) | 19 (78) |
| *Missing* | 1 (2.6) | 1 (9.1) | 2 (4) |
| **Household Income [n (%)]** |  |  |  |
| <499 Gourdes Haitian | 6 (15.4) | 0 (0) | 6 (12) |
| 500 – 2499 Gourdes Haitian | 14 (35.9) | 1 (9.1) | 15 (30) |
| 2500 – 4999 Gourdes Haitian | 7 (18) | 1 (9.1) | 8 (16) |
| >5000 Gourdes Haitian | 9 (23.1) | 6 (54.6) | 15 (30) |
| *Missing* | 3 (7.7) | 3 (27.3) | 6 (12) |
| **Average Number Dependents [mean (SD)]** | 3.58 (2.9) | 4.0 (3.7) | 3.66 (3) |
| **Substance Use** |  |  |  |
| Number who Smoke Cigarettes [n(%)] | 16 (41.0) | 5 (45.5) | 21 (42) |
| Number who Smoke Marijuana [n(%)] | 8 (20.5) | 3 (27.3) | 11 (22) |
| *Missing* | 0 (0) | 1 (9.1) | 1 (2.0) |
| **Number of Hours worked prior to**<br>**Incarceration [median (IQR)]** | 22.5 (8-48) | 10.0 (4-78) | 11.0 (4-78) |
| *Missing* | 3 (27.3) | 5 (12.8) | 8 (16) |
| **Alcohol Use** |  |  |  |
| Number who Use Alcohol [n(%)] | 21 (53.85) | 7 (63.64) | 28 (56) |
| Number who Drink Alcohol Daily<br>(n[%]) | 10 (47.62) | 4 (57.14) | 14 (50) |
| **ZLDSI Depression Scale** |  |  |  |
| Average Score [mean (SD)] | 13.85 (8.1) | 14.27 (8.74) | 13.9 (8.17) |
| Considered Depressed * [n(%)] | 25(64.10) | 8(72.73) | 33 (66) |
| **Stigma Score** |  |  |  |
| Internal Perceptions and Emotions | 1.67 (2.30) | 1.73 (1.49) | 1.68 (2.13) |
| Disclosure | 3.44 (4.36) | 4.00 (2.53) | 3.56 (4.01) |
| External Perceptions | 2.00 (3.16) | 2.91 (3.21) | 2.20 (3.16) |
| External Actions | 2.33 (3.52) | 3.27 (3.07) | 2.54 (3.42) |
| Courtesy Stigma | 0.26 (0.82) | 0.27 (0.91) | 0.26 (0.83) |
| *Total [mean (SD)]* | 9.69 (12.4) | 12.2 (7.41) | 10.2 (11.4) |

\* Cut-off score for depression on the ZLDSI scale was a score of 13 or higher.
\*\* Missing data due to participants leaving questions unanswered or reporting “Don’t Know” on survey.

As for the ZLDSI depression scale, the average score for those in VDOT sites was 13.85 (SD=8.1) with 25/39 (64.10%) participants considered depressed due to a score of 13 or greater. The average score was a 14.27 (SD=8.74) among DOT sites, and 8/11 (72.73%) of participants in this control prison met criteria for depression.

Results from the Cronbach’s alpha assessment are shown in *Table 2*. The inter-item reliability for the ZLDSI scale was high (13 items; a 0.77), signifying that no single item on the scale could be taken out to increase validity. The Cronbach’s alpha for the stigma scale was 0.68 (15 items), which did not show significant internal validity among the items for this study population, signifying that individual questions should not be analyzed separately to represent stigma as characterized in the original study.

**Table 2.** Cronbach Alpha for ZLDSI Depression Scale and Stigma Scale: Incarcerated TB patients, Haiti, 2019- 2020.

| Cronbach Alpha Value |  |  |  |  |  |  |  |
| --- | --- | --- | --- | --- | --- | --- | --- |
| Instrument Name | Description | Interpretation | Items | Number of subjects n | Literature alpha | Observed alpha | Response Range |
| **ZLDSI Depression Scale** | Measures participant's level of depression | A higher score indicates more mental distress congruent with depression<br>Instrument range: (0-3) | 13 | 50 | 0.89* | 0.77 | (0-35) |
| **Stigma Scale** | Measures the level of stigma that a participant is experiencing | A higher mean score indicates that a participant is experiencing higher levels of stigma<br>Instrument range: (0-3) | 20 in Haitian Stigma Scale ** (15 were used for this analysis) | 50 | > 0.80 ** | 0.68 | (0-35) |

\* Rasmussen *et al.* (2014)
\*\* Coreil *et al.* (2010)

Adherence to TB regimen was reported as 100% while persons resided in the control prisons that used DOT. In the intervention prisons, all subjects received VDOT while incarcerated; one participant who was surveyed left before ending treatment and completed his medication course in the community. If videos could not be made for technical reasons, correctional officers reported to the study office if they observed doses taken. When factoring in non-video recorded doses observed by correctional officers, the adherence of persons while residing in intervention prisons was >99%, but in the project overall, video-recorded adherence of doses was 80%, per SureAdhere logs (de Groot et al., 2022).

### Logistic Regression for Depression

Depression symptomatology was coded as a binary variable for this part of the analysis, where a score of 13 or higher on the ZLDSI score signified the manifestation of depression. From henceforth in this analysis, we will refer to a score equal to or greater than 13 on the screening scale for depression as *depression*.

An area of interest was the association between substance use (cigarette use, marijuana use, and alcohol consumption) and depression. (*Table 3*) After conducting univariate analyses with these variables, alcohol consumption showed the strongest association with depression, so it was left in the final model. Cigarette use and marijuana use were excluded from the final model due to their reduction of the final model’s significance. Based on previous literature studying factors affecting depression, we kept age and income in the model. Education trended towards significance, so this variable was also kept in the model.

**Table 3.** Unadjusted odds ratio and odds ratio adjusted for age, education, and income for factors associated with depression;^a^ Incarcerated TB patients, Haiti, 2019-2020.

|  | P-Value | Unadjusted OR | 95% CI | P-Value | Adjusted OR | 95% CI |
| --- | --- | --- | --- | --- | --- | --- |
| **Marijuana Use** | 0.077 | 6.96 | (0.81, 59.84) | n/a | n/a | n/a |
| **Cigarette Use** | **0.018** | 5.60 | (1.35, 23.23) | n/a | n/a | n/a |
| **Alcohol Consumption** | **0.008** | 5.52 | (1.53, 19.86) | **0.027** | **8.02** | **(1.26, 50.89)** |
| **Age** |  |  |  |  |  |  |
| **Below 31** |  | Reference |  |  |  |  |
| **31 and Above** | 0.209 | 0.44 | (0.12, 1.58) | **0.037** | **0.12** | **(0.02, 0.88)** |
| **Education** |  |  |  |  |  |  |
| **No education** |  | Reference |  |  |  |  |
| **Some level of education** | 0.078 | 3.00 | (0.89,10.54) | 0.123 | 3.84 | (0.69, 21.28) |
| **Financial income** |  |  |  |  |  |  |
| **Below 2500 Gourdes Haitian** |  | Reference |  |  |  |  |
| **2500 and more Gourdes Haitian** | 0.277 | 0.49 | (0.13, 1.78) | 0.622 | 0.66 | (0.13, 3.45) |
| **First time incarcerated** | 0.746 | 1.6 | (0.06, 42.54) | n/a | n/a | n/a |
| **Site** |  |  |  |  |  |  |
| **DOT sites** |  | Reference |  |  |  |  |
| **VDOT sites** | 0.595 | 0.67 | (0.13, 2.75) | n/a | n/a | n/a |
| **Stigma<sup>b</sup>** |  |  |  |  |  |  |
| **Less than score of 10** |  | Reference |  |  |  |  |
| **Score of 10 or higher** | 0.064 | 3.26 | (0.97, 12.27) | n/a | n/a | n/a |
| **Number of Dependents** |  |  |  |  |  |  |
| **Less than 3** |  | Reference |  |  |  |  |
| **3 or greater** | 0.739 | 1.23 | (0.35, 4.21) | n/a | n/a | n/a |
| **Number of hours worked prior to incarceration** |  |  |  |  |  |  |
| **Less than 10** |  | Reference |  |  |  |  |
| **10 or more** | 0.113 | 2.91 | (0.80, 11.65) | n/a | n/a | n/a |

<sup>a</sup> Depression is denoted by a score of 13 or higher on the ZLDSI depression scale
<sup>b</sup> The Courtesy Stigma level was omitted due to only having one question.

The adjusted model examined associations between depression and alcohol use, adjusting for age, education, and income. As the participant’s age increased to 31 years and older, the probability of experiencing depression decreased. Higher income trended towards being protective against depression in this model, although the association was not statistically significant (95% CI 0.13, 3.45).. When looking at education, those who had some years of education were 3.84 times more likely to have a positive score for depression compared to participants with no education; however, this relationship was not statistically significant (95% CI 0.69, 21.28). When studying interaction terms, there were no significant terms found in this model.

### Analysis of Substance Use

Baseline characteristics of those who reported substance and alcohol use are noted in *Table 4.* When observing alcohol use as a dependent variable, we found that cigarette use was significantly associated with alcohol use, controlling for depression, age, financial income, and education (95% CI 2.54-337.68).

**Table 4.** Study Characteristics of Participants with Substance Use; Incarcerated TB patients.

Haiti, 2019-2020*
|  | **Marijuana Use** | **Cigarettes Use** | **Alcohol Consumption** |
| --- | --- | --- | --- |
| All participants (n[%]) | 11 (100) | 21 (100) | 28 (100) |
| Age (mean[SD]) | 27.3 (4.9) | 30.4 (8.8) | 30.2 (8.6) |
| One or more years of education | 7 (63.6) | 16 (76.2) | 19 (67.9) |
| Average years of Education | 3.50 (4.1) | 5.16 (4.3) | 4.56 (4.3) |
| Financial Income n[%] |  |  |  |
| <499 Gourdes Haitian | 1(0) | 0 (0) | 2 (7.1) |
| 500 – 2499 Gourdes Haitian | 2 (18.2) | 5 (23.8) | 6 (21.4) |
| 2500 – 4999 Gourdes Haitian | 2 (18.2) | 5 (23.8) | 5 (17.9) |
| >5000 Gourdes Haitian | 7 (63.6) | 11 (52.4) | 12 (42.9) |
| Average Depression (mean[SD]) | 16.2 (4.4) | 17.3 (6.5) | 16.1 (6) |
| Depressed (13+ score) | 10 (90.9) | 18 (85.7) | 23 (82.1) |
| Stigma, Total (mean[SD]) | 10.5 (10.8) | 11.4 (12.9) | 10.2 (10.9) |
| Internal Perceptions and Emotions | 1.73 (2) | 1.86 (2.3) | 1.57 (2.03) |
| Disclosure | 3.00 (3.2) | 3.33 (4) | 3.43 (3.8) |
| External Perceptions | 2.55 (3.3) | 2.67 (3.2) | 2.18 (2.9) |
| External Actions | 2.73 (3.3) | 2.95 (3.8) | 2.64 (3.3) |
| Courtesy Stigma | 0.55 (1.2) | 0.62 (1.2) | 0.36 (1) |
| First time incarcerated (n[%]) | 9 (81.8) | 15 (71.4) | 20 (71.4) |
| Smoke Marijuana (n[%]) | - | 9 (42.9) | 10 (35.7) |
| Smoke Cigarettes (n[%]) | 9 (81.8) | - | 19 (67.9) |
| Drink Alcohol (n[%]) | 10 (90.9) | 19 (90.5) | - |

## Discussion

In a Haitian prison population, a screening instrument administered in a cross-sectional survey of enrollees in a TB treatment project found depression in the majority of persons. Adherence to TB medication was reported to be essentially 100%. We examined the likelihood of a positive screen for depression as a secondary analysis where we analyzed its associations with substance use (cigarette use, marijuana use, and alcohol consumption). The study population mainly consisted of participants from VDOT sites (78%) as there was just one DOT control site surveyed. Levels of depression symptomatology did not differ across prisons by site, despite the literature showing that the experience of prison populations can be highly variable and site-specific (Cooley, 2019).

Cigarette use prior to incarceration was high, as has been observed elsewhere (Spaulding et al., 2018). Tobacco use was more prevalent in the prison population (42%) compared to marijuana (22%). This can be attributed to the fact that marijuana is illegal in Haiti (CIA, 2021). As a result, marijuana is available only for those who have the means to obtain it and believe they can evade authorities. As mentioned in the background, the majority of those who are incarcerated come from low-income households (Walmsley, 2016), though in this specific population we saw roughly equal amounts of people coming from low-income and middle-income households. Out of the 11 people who smoke marijuana in this population, 7 (64%) made more than 5000 Gourdes Haitian a month, showing that individuals with higher income tend to smoke more marijuana compared to those with lower income *Table 4*.

Alcohol use was also highly prevalent in this population, as was expected (Yi et al., 2017). More than half of participants reporting that they drank alcohol (54% in VDOT prisons and 64% in DOT prisons). Historically Haiti was one of the top liquor-producing countries in the world. Their traditional drink is a rum known as clairin (Feingold and Weinstein, 2020). With its production has come a very rich culture around drinking alcohol in Haiti, and this could also contribute to alcohol use being one of the biggest substances of concern in the country (Angulo- Arreola et al., 2011).

For the ZLDSI depression scale, mean scores for both VDOT and DOT prison sites, 13.85 and 14.27 respectively, were greater than the depression cut-off score of 13, signifying that the average person living with TB and incarcerated in these prisons was depressed. Interestingly, the mean ZLDSI depression score for VDOT and DOT prison sites were similar, signifying no significant differences in depression between the two sites. Our analysis sought to find associations of different variables with depression among this population. As seen in our study, we found significant associations within the bivariate analyses between cigarette use and alcohol consumption, with alcohol consumption having the largest association with depression (*Table 4*). Stigma and number of dependents were variables that trended towards significance during this analysis.

Along with alcohol consumption, we also kept age, education, and financial income in the final model. Previous research has shown significant associations between substance use and depression (Feingold and Weinstein, 2020), but these studies included a much larger study population that was not incarcerated or made up of people who were positive with TB. Studies in high income countries have reported a higher prevalence of depressed individuals in incarcerated populations compared to the general population (Berzofsky and Bronson, 2017).

Many of these associations yielded very large confidence intervals as would be expected in a small study. This could signify an instability problem where there may not have been complete separation of all participants who were significantly different from null values. Additionally, there was a significant percentage of missing values within this dataset and we did not perform analyses to impute missing data. Along with missing values were many values that were 0, further contributing to the “complete separation” problem. A future study could study the ZLDSI scale as a continuous variable where the focus would be more on the symptom severity for depression in incarcerated populations compared to an urge for further mental health treatment and care.

When observing alcohol use as a dependent variable, we found that cigarette use was significantly associated with alcohol use, controlling for depression, age, financial income, and education, however the findings were not very precise; the confidence interval was wide (95% CI 2.54-337.68). When observing marijuana use as a dependent variable, alcohol was significantly associated with marijuana use, controlling for age, financial income, first time incarcerated, and education level. Past studies have observed a similar relationship between perceived stress and depression in substance use treatment (McHugh et al., 2020).

The ZLDSI scale held internal consistency in our study with a Cronbach alpha value of a 0.77 (*Table II)*. When analyzing the Cronbach alpha value for the stigma scale, we found that our value was slightly different and did not hold as much internal validity as that in the literature. With the Cronbach alpha value of the stigma scale overall as 0.68 for this study, we found that the modified stigma scale did not perform well in this study. This was a substantial difference from the greater than 0.8 value found in literature (Coreil et al., 2010). Our modifications could have compromised the internal validity of our scale, leading to lower, unacceptable Cronbach alpha values.

Our research sought to find if an association between substance use and depression, demonstrated in US populations, was also present in the Haitian prison population. In this setting, where a diagnosis of depression is rare, we wanted to observe whether substance use could be used as an indicator for symptoms of depression. If that association were to be established, prison providers might first screen persons for substance use and then focus screening efforts for depression on persons affirming substance use disorder, in order to locate those who would benefit from treatment of depression.

### Limitations

This study held several limitations. The ZLDSI scale is not meant to replace the clinical diagnosis of depression by a licensed clinician. It is meant to be an indicator tool to encourage further screening and care for those who score higher on the ZLDSI scale.

For those who had already been incarcerated for years, it may have been a while since the time that people have used alcohol and other substances. Self-reported measures may have been inaccurate due to recall bias. Cross-sectional analysis diminishes our ability of establishing temporality in this analysis. With a longitudinal analysis, more information could be collected on the temporality of depression and substance use onset while also considering other factors such as site, age, education level, household income, number of visitors (as a proxy for social support), and number of dependents. Furthermore, we examined these associations in all-male prisons in Haiti, a country where nearly 97% of incarcerated persons are male (www.prisonstudies.org). Future studies can study associations within women’s prisons in the country. Lastly, study of depression among persons without tuberculosis would be helpful.

With a larger pool of participants, and a longitudinal study design, we could examine the effect of VDOT versus DOT on substance use, depression, and continuity of care. We examined these associations in all-male prisons. Future studies can study associations within women’s prisons. Additionally, due to the research staff’s limited ability to reach each site repeatedly, the number of surveys attained was limited. More research is needed to assess the accuracy of the ZLDSI scale within not only adults across the country, but people in Haiti’s correctional system.

Selection of the sites for VDOT was not random; it was implemented in the prisons located in remote areas of Haiti that are difficult for healthcare workers to visit regularly. While this study was being conducted, political unrest was prevalent throughout Haiti. An unexpected consequence was that the VDOT permitted continuation of TB therapy that otherwise would have stopped. With no present-functioning Haitian legislature and increased gang-related crime rates in the country, the effect of the unrest on mental health in Haiti remains unclear. The COVID-19 pandemic brought even more health challenges to Haiti’s prisons.

## Conclusion

The ZLDSI scale showed good internal validity for this study population of incarcerated individuals in Haitian prisons. This preliminary study should provide a basis for partnerships with organizations focused on delivering mental health care to resource-poor areas.

Measuring depression by the ZLDSI scale among participants displayed significant associations with prior substance use; however, more analyses need to be performed to confirm a strong and significant relationship between these variables. Education was seen as a significant modifier of the association between alcohol consumption and depression.

With mental health illness being an issue on a global scale, more research is needed around designing ethnoculturally appropriate mental health measurement tools, such as the ZLDSI scale for Haiti, to better assess mental health in low-resourced regions of the world. Applying appropriate scales to incarcerated populations, developed in the community from which incarcerated persons come, especially when proxy measurements indicate a high likelihood of depression, should be a high priority for the well-being of those in custody everywhere.

## Data Availability

All data produced in the present work are contained in the manuscript.

## Acknowledgements

We thank Dr. John May for his initial vision for the Correctional officer facilitated VDOT for TB treatment in Haitian Correctional Facilities project; Dr. Kelly Collins for her help with the SureAdhere platform; Dr. Andrew Rasmussen for important feedback about use of the Zanmi-Lasante Depression Symptom Inventory; and Alexandra Kauffman for help in preparing this manuscript for submission. We appreciate the support of the TB Research Advancement Center (TRAC, P30AI168386) of Emory/Georgia.

## Abbreviations used

DOT: direct observed therapy
TB: tuberculosis
VDOT: video direct observed therapy
ZLDSI: Zanmi-Lasante Depression Symptom Inventory

## Appendix

### Supplementary Tables

**Table S1:** English translation ZLDSI scale from Rasmussen et al. (2014) paper.

| Item | Depressed mood | Vegetative symptoms |
| --- | --- | --- |
| Thinking too much | .702 | — |
| Feeling down, discouraged or totally hopeless | .682 | — |
| Thoughts that you would be better off dead, or of hurting yourself in some way | .682 | — |
| Feeling you are a failure or feeling bad about yourself | .666 | — |
| Feeling you've lost the taste for doing anything | .653 | — |
| Feeling you have a constricted heart | .639 | — |
| Having no appetite | .527 | .452 |
| Crying or feeling like crying | .511 | — |
| Feeling tired or having little energy | .477 | .508 |
| Low energy | .430 | .468 |
| Moving or speaking so slowly that people have noticed | — | .626 |
| Difficulty sleeping without waking early | — | .861 |
| Difficulty falling asleep | — | .840 |

**Table S2:** Original ZLDSI scale from Partners in Health in Haitian Creole (Partners in Health Curriculum Toolkit, 2016)

|  | Pandan 15 jou ki sòt pase la yo, konbyen fwa yon nan pwoblèm sa yo te fatige ou ? | Di tou | Konbyen fwa yon nan pwoblèm sa yo te fatige ou ? | Pandan kèk jou (1–5 jou) | Plis pase yon semèn (6–9 jou) | Preske chak jou (10–15 jou) |
| --- | --- | --- | --- | --- | --- | --- |
| 1 | Santi ou de la la. | 0 | — | 1 | 2 | 3 |
| 2 | Santi kè sere. | 0 | — | 1 | 2 | 3 |
| 3 | Kalkile twòp. | 0 | — | 1 | 2 | 3 |
| 4 | Kriye oubyen anvi kriye | 0 | — | 1 | 2 | 3 |
| 5 | Santi anyen preske pa enterese ou. | 0 | — | 1 | 2 | 3 |
| 6 | Santi ou kagou. dekouraje ak lavi, oubyen pèdi espwa nèt ale. | 0 | — | 1 | 2 | 3 |
| 7 | Gen difikilte pou dòmi pran ou. | 0 | — | 1 | 2 | 3 |
| 8 | Santi ou fatige oubyen ou manke fòs. | 0 | — | 1 | 2 | 3 |
| 9 | Ou pa gen apeti. | 0 | — | 1 | 2 | 3 |
| 10 | Ou santi lavi-w pase mal oubyen ou santi-w pa alèz ak tèt-w. | 0 | — | 1 | 2 | 3 |
| 11 | Fè mouvman oubyen pale tèlman dousman, menm lòt moun wè sa. | 0 | — | 1 | 2 | 3 |
| 12 | Ou di nan tèt ou: Pito-w te mouri, oubyen ou gen lide pou fè tèt-w mal. | 0 | — | 1 | 2 | 3 |
| 13 | Gen difikilte pou rete dòmi jouk li jou. | 0 | — | 1 | 2 | 3 |
| **Totals** |  |  |  | **(+)** | **(+)** |  |

(=) **ZLDSI Score** \_\_\_\_\_

**Table S3:**
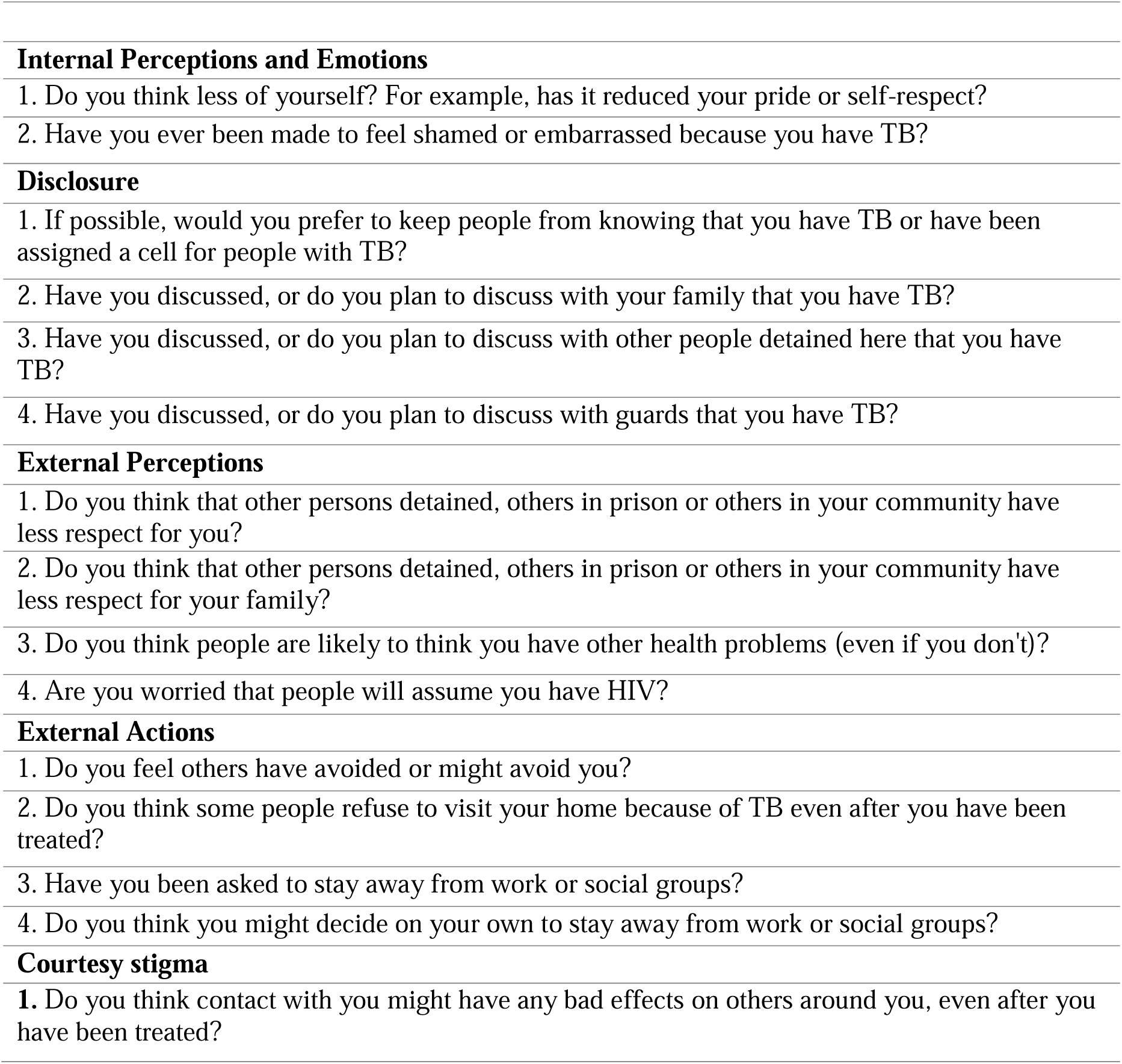
Stigma scale used for this study. Modified from Coreil et al. *(2010)*

|  |
| --- |
| **Internal Perceptions and Emotions** |
| 1. Do you think less of yourself? For example, has it reduced your pride or self-respect? |
| 2. Have you ever been made to feel shamed or embarrassed because you have TB? |
| **Disclosure** |
| 1. If possible, would you prefer to keep people from knowing that you have TB or have been assigned a cell for people with TB? |
| 2. Have you discussed, or do you plan to discuss with your family that you have TB? |
| 3. Have you discussed, or do you plan to discuss with other people detained here that you have TB? |
| 4. Have you discussed, or do you plan to discuss with guards that you have TB? |
| **External Perceptions** |
| 1. Do you think that other persons detained, others in prison or others in your community have less respect for you? |
| 2. Do you think that other persons detained, others in prison or others in your community have less respect for your family? |
| 3. Do you think people are likely to think you have other health problems (even if you don't)? |
| 4. Are you worried that people will assume you have HIV? |
| **External Actions** |
| 1. Do you feel others have avoided or might avoid you? |
| 2. Do you think some people refuse to visit your home because of TB even after you have been treated? |
| 3. Have you been asked to stay away from work or social groups? |
| 4. Do you think you might decide on your own to stay away from work or social groups? |
| **Courtesy stigma** |
| 1. Do you think contact with you might have any bad effects on others around you, even after you have been treated? |

**Table S4:** Original Full stigma scale from Coreil et al. *(2010) paper*.

| Factor Label and Scale Items | Item abbreviation |
| --- | --- |
| *Internal perceptions and emotions.* |  |
| Would Jean think less of himself because he has TB? Might he have less pride or self-respect? | Think less of self |
| Do you think Jean might feel ashamed or embarrassed because of this problem? | Feel ashamed |
| Would Jean be worried that he might have to spend time in a TB hospital/sanatorium? | Worry about sanatorium |
| *Disclosure* |  |
| Do you think Jean would prefer to keep others from knowing about this problem? | Hide from others |
| Do you think that Jean would discuss this problem with family members? | Disclose to family |
| Do you think that Jean would discuss this problem with close friends? | Disclose to friends |
| Do you think that Jean would discuss this problem with neighbors? | Disclose to neighbors |
| Do you think that Jean would discuss this problem with church members? | Disclose to church members |
| Do you think that Jean would discuss this problem with co-workers? | Disclose to co-workers |
| *External perceptions* |  |
| If others in the community came to know about it, would they have less respect for Jean? | Loss of respect |
| Would others believe that Jean is dangerous because of this condition? | Think dangerous |
| Even if Jean didn't have any other health problems, are people likely to think he does because he has TB? | Assume has other conditions |
| Would people assume he has HIV? | Assume has HIV |
| *External actions* |  |
| Do you think people might say or do anything to hurt Jean because of his illness? | Others hurtful |
| Do you think others might avoid Jean because of this problem? | Others avoid |
| Would other people refuse to visit Jean's home because of this condition even after treatment? | Others refuse to visit |
| Would this condition cause problems for Jean at work if people knew about it? | Problems at work |
| Do you think that Jean's wife will remain with him and be supportive over the entire course of the treatment? | Wife supportive |
| If Jean were unmarried, do you feel that even after having received treatment and having been cured, Jean might still have difficulty getting married, as a result of his illness? | Difficulty marrying |
| *Courtesy stigma* |  |
| If others in the community came to know about it, would they have less respect for Jean's family? | Less respect for family |
| If he had children, do you think the problem might cause social problems for Jean's children in the community? | Problems for children |
| Would contact with Jean have bad effects on others around him even after he is treated? | Effects on others |
| *Haitian identity* |  |
| Is it more embarrassing for Jean to have TB because he is Haitian than it would be for other people in Florida? | Embarrassing because Haitian |
| Because Jean is Haitian, will people think worse of him than others with TB? | Think worse because Haitian |

**Table S5:**
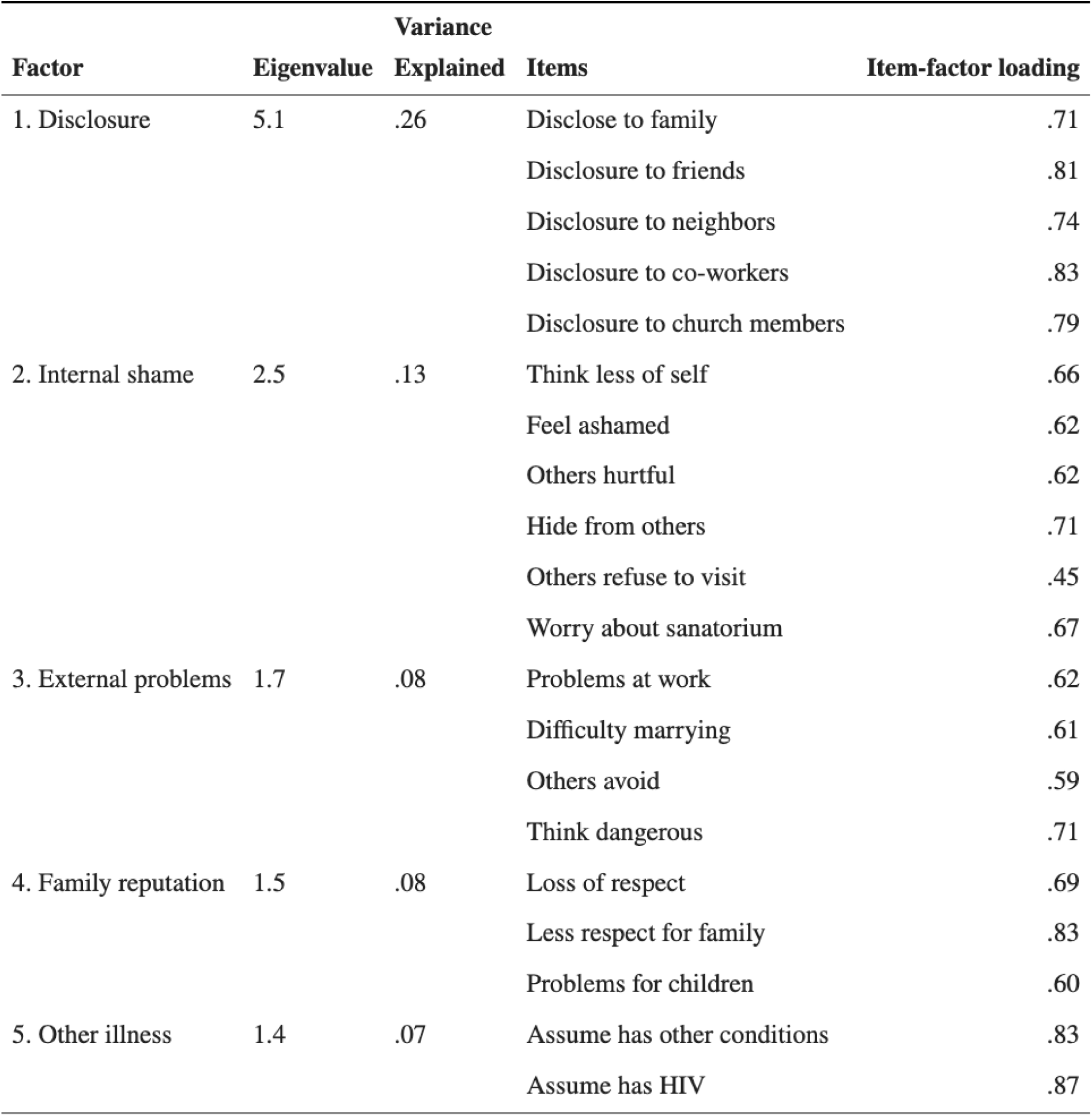
Original stigma scale from Coreil et al. *(2010) paper that was formulated for their Haitian community*.

## References

Berzofsky, M. Dr.PH, & Bronson, J, PhD. (2017). Indicators of Mental Health Problems Reported by Prisoners and Jail Inmates, 2011-12. US Bureau of Justice Statistics. Washington, DC. https://bjs.ojp.gov/content/pub/pdf/imhprpji1112.pdf

Cénat, J. M., McIntee, S. E., & Blais-Rochette, C. (2020). Symptoms of posttraumatic stress disorder, depression, anxiety and other mental health problems following the 2010 earthquake in Haiti: A systematic review and meta-analysis. Journal of Affective Disorders, 273, 55–85. 10.1016/j.jad.2020.04.046

Charles, M., Richard, M., Joseph, P., Bury, M. R., Perrin, G., Louis, F. J., Fitter, D. L., Marston, B. J., Deyde, V., Boncy, J., Morose, W., Pape, J. W., & Lowrance, D. W. (2017). Trends in Tuberculosis Case Notification and Treatment Success, Haiti, 2010–2015. The American Journal of Tropical Medicine and Hygiene, 97(4_Suppl), 49–56. 10.4269/ajtmh.16-0863

Cooley, C. (2019). Escaping the Prison of Mind: Meditation as Violence Prevention for the Incarcerated. Health Promotion Practice, 20(6), 798–800. 10.1177/1524839919869924

Coreil, J., Mayard, G., Simpson, K. M., Lauzardo, M., Zhu, Y., & Weiss, M. (2010). Structural forces and the production of TB-related stigma among Haitians in two contexts. Social Science & Medicine, 71(8), 1409–1417. 10.1016/j.socscimed.2010.07.017

de Groot, L. M., Straetemans, M., Maraba, N., Jennings, L., Gler, M. T., Marcelo, D., Mekoro, M., Steenkamp, P., Gavioli, R., Spaulding, A., Prophete, E., Bury, M., Banu, S., Sultana, S., Onjare, B., Efo, E., Alacapa, J., Levy, J., Morales, M. L. L., & Bakker, M. I. (2022). Time Trend Analysis of Tuberculosis Treatment While Using Digital Adherence Technologies—An Individual Patient Data Meta-Analysis of Eleven Projects across Ten High Tuberculosis-Burden Countries. Tropical Medicine and Infectious Disease, 7(5), 65. 10.3390/tropicalmed7050065

Feingold, D., & Weinstein, A. (2020). Cannabis and Depression. Cannabinoids and Neuropsychiatric Disorders, 67–80. 10.1007/978-3-030-57369-0_5 Get Ready to Fall in Love with Haiti’s One-of-a-Kind Rum. (2021, January 25). Liquor. https://www.liquor.com/articles/clairin-rum/

Garfein, R. S., Collins, K., Muñoz, F., Moser, K., Cerecer-Callu, P., Raab, F., Rios, P., Flick, A., Zúñiga, M. L., Cuevas-Mota, J., Liang, K., Rangel, G., Burgos, J. L., Rodwell, T. C., & Patrick, K. (2015). Feasibility of tuberculosis treatment monitoring by video directly observed therapy: a binational pilot study. The International Journal of Tuberculosis and Lung Disease, 19(9), 1057–1064. 10.5588/ijtld.14.0923

Garfein RS, Liu L, Cuevas-Mota J, Collins K, Muñoz F, Catanzaro DG, Moser K, Higashi J, Al-Samarrai T, Kriner P, Vaishampayan J. Tuberculosis treatment monitoring by video directly observed therapy in 5 health districts, California, USA. Emerging infectious diseases. 2018 Oct;24(10):1806.

*Haiti | World Prison Brief*. (2021). World Prison Brief. https://www.prisonstudies.org/country/haiti

Health Through Walls website (2021). https://www.healththroughwalls.org/haiti.htm Last accessed 2 August 2022.

Hill, K., Wainwright, V., Stevenson, C., Senior, J., Robinson, C., & Shaw, J. (2022). Prevalence of mental health and suicide risk in prisons in low- and middle-income countries: a rapid review. The Journal of Forensic Psychiatry & Psychology, 33(1), 37–52. 10.1080/14789949.2021.2016891

Legha RK, Gerbasi ME, Smith Fawzi MC, Eustache E, Therosme T, Fils-Aime JR, Raviola GJ, Affricot E, Pierre EL, Alcindor Y, Severe J, Boyd KA, Grelotti DJ, Darghouth S, Rasmussen A, Becker AE. A validation study of the Zanmi Lasante Depression Symptom Inventory (ZLDSI) in a school-based study population of transitional age youth in Haiti. Confl Health. 2020 Feb 28;14:13. doi: 10.1186/s13031-020-0250-9. PMID: 32140176; PMCID: PMC7048134.

McHugh, R. K., Sugarman, D. E., Meyer, L., Fitzmaurice, G. M., & Greenfield, S. F. (2020). The relationship between perceived stress and depression in substance use disorder treatment. Drug and Alcohol Dependence, 207, 107819. 10.1016/j.drugalcdep.2019.107819

Partners in Health Curriculum Toolkit (2016) Mental Health Innovation Network. Available at http://www.mhinnovation.net/resources/partners-health-curriculum-toolkit (Accessed 23 January 2022).

Rasmussen, A., Eustache, E., Raviola, G., Kaiser, B., Grelotti, D. J., & Belkin, G. S. (2014). Development and validation of a Haitian Creole screening instrument for depression. Transcultural Psychiatry, 52(1), 33–57. 10.1177/1363461514543546

Spaulding AC, Eldridge GD, Chico CE, Morisseau N, Drobeniuc A, Fils-Aime R, Day C, Hopkins R, Jin X, Chen J, Dolan KA (2018). Smoking in correctional settings worldwide: prevalence, bans, and interventions. Epidemiologic reviews. 2018 Jun 1;40(1):82-95.

Walmsley, R. (2016). World Prison Population List. Prison Studies. https://www.prisonstudies.org/sites/default/files/resources/downloads/world_prison_population_list_11t h_edition_0.pdf

The World Factbook 2021. Washington, DC: Central Intelligence Agency, 2021. https://www.cia.gov/the-world-factbook/

Yi, Y., Turney, K., & Wildeman, C. (2017). Mental Health Among Jail and Prison Inmates. American journal of men’s health, 11(4), 900–909. 10.1177/1557988316681339

